# An Exploratory Stability Selection (ESS) Framework for Robust Predictor Discovery: An Application to Physical Resilience in Aging Populations

**DOI:** 10.64898/2026.08.01.26359469

**Authors:** Marissa C. Ashner, Virginia B. Kraus, Heather E. Whitson, Corey B. Simon, Janet L. Huebner, Akshay Bareja, Chelsea R. Perfect, Laura Pietrosimone, Katherine S. Hall, Cathleen S. Colón-Emeric, Sarah B. Peskoe

## Abstract

Identifying biological and clinical signals that consistently predict physical resilience, defined as one’s ability to maintain or regain function following a health stressor, is essential for advancing precision approaches to aging and recovery. High-dimensional datasets hold tremendous promise but pose analytic challenges due to correlation, distributed signals, instability, and sensitivity to analytic choices. The complexity of these data requires strategies that prioritize transparency and stability in variable selection. We present a resampling-based statistical framework, the Exploratory Stability Selection (ESS) framework, designed for hypothesis-generating predictor discovery. ESS is an ensemble-style variable selection technique that integrates multiple resampling strategies and sparsity levels, enabling exploration of robustness and context-dependence across diverse data perturbations.

We demonstrate the utilization of the ESS framework with a data example using the PRIME-KNEE study, which examines physical resilience in older adults undergoing elective total knee arthroplasty. ESS analyses were applied to clinical-only, plasma biomarker-only, and combined predictor sets to evaluate the stability and competitiveness of candidate variables associated with the probability of having a highly resilient recovery trajectory for pain interference. The data example highlights how ESS distinguishes highly stable predictors from context-dependent signals whose selection varies with predictor competition and analytic configuration. ESS retains configuration-level results and summarizes stability metrics across configurations to provide insight into the subsequent prioritization and validation of candidate predictors. This framework is well-suited for hypothesis-generating variable selection problems common to exploratory resilience research and other aging-related applications that involve complex, multi-domain predictor sets.

## 1. Introduction

As the aging population continues to grow, there has been an increasing emphasis on resilience-focused research in older adults. In fact, the National Institute on Aging (NIA) has convened several workshops to discuss the concept of resilience in aging over the last decade (Hadley et al 2017, Abadir et al 2023, Colón-Emeric et al 2024). Resilience in older adults refers to the ability to resist or recover from a stressor (Windle 2011, Hadley et al 2017) and can span multiple domains, including cognitive, psychosocial, and physical resilience. While the motivating application in this work focuses on physical resilience, the proposed framework is more broadly applicable across domains and composite operationalizations or measures (e.g., frailty or quality of life). Physical resilience, defined as a person’s ability to maintain or regain function after a physical health stressor, has become a priority in geroscience (Abadir et al 2023, Walston et al 2023, Grigoras et al 2025, Xu et al 2025). Physical resilience is a dynamic, multi-faceted construct, underscoring the need for a deeper understanding of its intrinsic and extrinsic influences in order to inform appropriate interventions and more broadly understand healthy aging capacity (Windle 2011, Whitson et al 2016, Hadley et al 2017, Grigoras et al 2025). While many studies have identified a range of factors influencing psychologically-measured resilience, these results are subject to between-study heterogeneity and do not necessarily extend to physical resilience (Górska et al, 2022).

Several recently-completed cohort and electronic health record-based studies have collected an abundance of data from multiple sources before and after elective physical stressors to gain insight into the factors that influence physical resilience (Whitson et al 2021, Laskow et al 2022, Walston et al 2023, Xu et al 2025). Most of these studies integrate both clinical and biological data, creating opportunities to unveil interactions between these factors and resilience outcomes. However, the diversity, high-dimensionality, and correlated structure of these multi-domain predictor sets, combined with modest sample sizes, create analytic challenges. Thus far, analyses of these data have focused on quantifying associations between targeted clinical predictors and resilience outcomes (Laskow et al 2022, Varadhan et al 2024, Colón-Emeric et al 2026). Biomarker discovery in physical resilience, however, is comparatively understudied, despite the growing availability of biological data (Windle 2011, Walston et al 2023). Although earlier studies on biomarkers related to aging and physical resilience have helped guide the current biomarker collection process (Whitson et al 2021, Colón-Emeric et al 2023), the number of potential predictors is still very large. As a result, biomarker analyses of this nature and size should be considered exploratory and aimed at generating hypotheses, rather than testing specific ones. Therefore, while the eventual goal is to validate biomarkers for clinical translation, the immediate need is to pare down the number of candidate biomarkers for downstream confirmatory analyses in resilience research.

Variable selection (or feature selection) methods are often used in settings with high-dimensional predictor sets like those we encounter in physical resilience research. While many variable selection approaches exist, no single method performs optimally across analytic goals and data structures (Barbieri et al 2024, Wang et al 2019). Notably, there are three unique goals for the optimization of a variable selection method, including prediction, variable ranking, and recovery of the underlying active set (Wang et al 2019). In hypothesis-generating factor identification for physical resilience research, the primary goal is reliable recovery of the truly informative variables. LASSO (Least-Absolute Shrinkage and Selection Operator) is a commonly used and interpretable selection method, but it, as well as many other approaches, can be unstable, yielding results that are sensitive to perturbations of the data or choice of tuning parameters (Leng et al 2006, Budhraja et al 2023, Hédou et al 2024, Kissel and Mentch 2024).

Stability selection and Bolasso (Bootsrap-enhanced LASSO) are both variable selection techniques that pool active sets of variables across resampled subsets of the data (Bach 2008, Meinhausen and Buhlmann 2010, Shah and Samworth 2013). These approaches can be viewed as ensemble-style methods that have proven effective by leveraging data perturbation to overcome stability limitations that exist within single-model fits (Seijo-Pardo 2017, Budhraja et al 2023, Büyükkeçeci and Okur 2023). However, these methods remain sensitive to the choice of certain hyperparameters, and their distinct resampling strategies entail method-specific strengths and limitations (De Bin et al 2016).

In this paper, we introduce an Exploratory Stability Selection (ESS) framework, inspired by both classic stability selection and Bolasso but increases robustness to analytic perturbations by integrating multiple resampling strategies and sparsity levels within a single framework. ESS analyses are specifically designed for hypothesis-generating predictor discovery, particularly in physical resilience research where signals may appear weaker due to correlation and distribution across multiple predictor domains. Instead of performing formal inferences or building generalizable prediction models, ESS aims to characterize the underlying structures of the relationship between multi-domain candidate predictors and resilience. ESS retains stability metrics to transparently characterize the redundancy and competitiveness of predictors across different analytic perturbations, which will help guide prioritization of candidates for downstream validation.

The remainder of the paper is organized as following: Section 2 (Methods) describes the ESS architecture, components, execution, and recommended practices; Section 3 (Data Example) illustrates the framework using data from the PRIME-Knee study; Section 4 (Discussion) discusses the implications of the framework and results, along with limitations and potential downstream uses of ESS outputs.

## 2. Methods

### 2.1. Overview of the Exploratory Stability Selection (ESS) Framework

Our proposed ESS framework is a resampling-based process that characterizes the robustness of candidate predictors across multiple analytic perturbations. Unlike classic stability selection (Meinhausen and Buhlmann 2010, Shah and Samworth 2013), ESS is explicitly exploratory and does not aim to provide formal error-control guarantees. Instead, it aims to provide a very robust stable set of predictors by integrating multiple resampling strategies and sparsity levels, thereby capturing different types of sampling variation and sample perturbation and reducing the risk of instability driven by the LASSO tuning parameter. In hypothesis-generating variable selection, the key objective is to identify variables whose associations with the outcome are consistent, robust, and stable, thereby highlighting promising candidates for future study.

### 2.2. ESS Algorithm and Analytic Workflow

Here, we formally define the analytic workflow underlying ESS, which is summarized in Figure 1. This workflow is organized into three stages: inputs, configuration-level analysis, and cross-configuration summaries, shown from left to right in the diagram.

**Figure 1:**
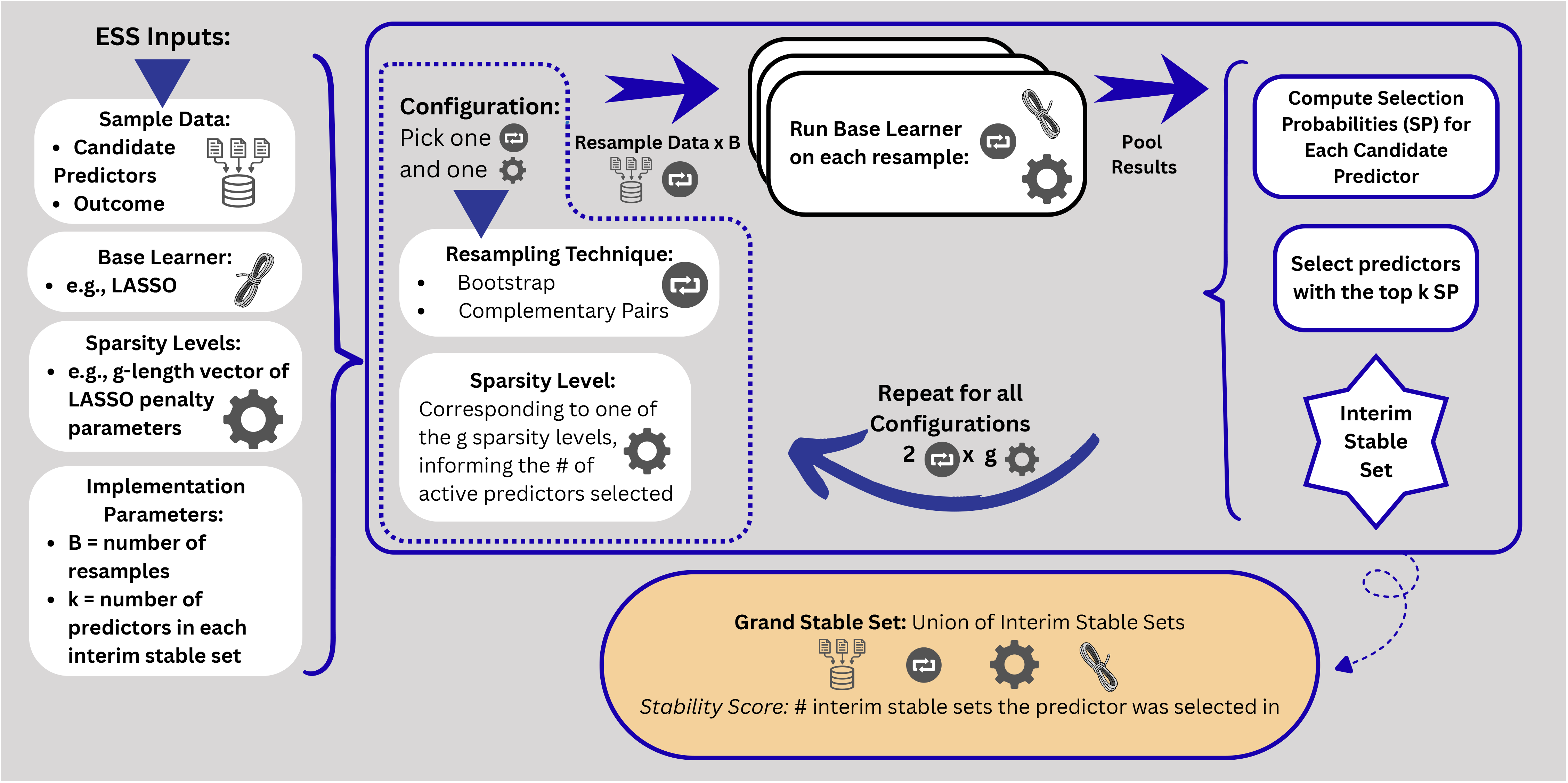
Diagram of the Exploratory Stability Selection (ESS) Framework

The ESS framework, where configurations of resampling and sparsity levels are used to compute selection probabilities and form interim stable sets that are ultimately combined into a grand stable set with stability scores.

First, ESS requires four key inputs, and recommendations for selecting these inputs are provided in Section 2.7.2:

1. *Sample Data* must include an outcome and a candidate predictor set, measured on the same individuals; in this application, we will use a continuous outcome
2. A *base learner* specifies the variable selection method applied within each model fit; in this application, we use LASSO
3. A set of *sparsity levels* defines the range of penalization values used to control the number of selected predictors, allowing exploration of different levels of model sparsity and predictor competition; the specific parameter associated with sparsity level may differ by choice of base learner
4. *Implementation parameters* include the number of resamples per configuration (*B*) and the size of the interim stable set (*k*)

Given these inputs, ESS constructs a set of analytic configurations using all combinations of resampling strategies (i.e., complementary pairs subsampling and bootstrap resampling) and sparsity levels.

For each analytic configuration, ESS applies the base learner to *B* resampled versions of the data. The repeated model fits are aggregated to generate configuration-specific selection probabilities for each candidate predictor, with *B* influencing the reliability of the estimated probabilities. Within each configuration, all predictors are then ranked by their selection probabilities, and the top-*k* predictors define the *interim stable set*. This process is repeated for all each analytic configuration, producing multiple interim stable sets, reflecting stability under different forms of data perturbation and levels of predictor competition.

Finally, the *grand stable set* is the union of all interim stable sets. ESS summarizes results using stability metrics, including selection probabilities and stability scores (i.e., configuration counts), along with descriptive effect estimates for predictors retained in the grand stable set. In the remainder of this section, we will describe the design choices, stability metrics, and practical considerations for implementation of ESS.

### 2.3. Structural Design Choices in ESS

#### 2.3.1. Resampling Strategies

One defining feature of ESS is the framework’s incorporation of multiple resampling strategies to capture different sources of sampling variability. We present two resampling techniques, complementary pairs subsampling and bootstrap resampling, which have complementary strengths and flaws. We acknowledge that ESS can be scalable to any number and type of resampling strategies, but we have found these two to allow us to strike a balance between interpretability, robustness, and comprehensiveness. Additionally, both resampling strategies discussed in this application are flexible and compatible with any base learner.

The first resampling strategy is complementary pairs (or CPSS), which originated as an extension of the original stability selection algorithm (Shah and Samworth 2013). The data are randomly split into two disjoint halves and each subsample is analyzed separately. To obtain *B* samples, we split the data *B*/2 times to retain *B* total subsamples. Since CPSS subsamples are structured half-sample perturbations of the data, each subsample is fit under a reduced effective sample size. As a result, this resampling structure has less tolerance for spurious or weak signals, since the candidate variables must survive perturbation with less information and random splitting of the data. These disjoint, non-overlapping subsamples improve upon random n/2 subsampling, which was proposed in the original stability selection paper (Meinhausen and Buhlmann 2010), because CPSS reduces dependence between subsamples (Shah and Samworth 2013). ESS uses CPSS primarily to explore robustness under this specific type of sampling variation, rather than to enforce the theoretical error-control guarantees that CPSS is typically associated with.

The second resampling strategy incorporated into ESS is the bootstrap. This is a common resampling strategy used in statistics and data science, recognized by non-statisticians as well. A bootstrap sample is created by sampling with replacement and full sample size, which captures variability under the empirical data-generating process. In contrast to CPSS, bootstrap resampling preserves total information in terms of effective sample size and reflects a larger range of sampling variability due to the omission and repetition of certain observations. Therefore, bootstrap is useful to include in ESS as a complementary resampling strategy to CPSS. This can, however, inflate some selection frequencies by oversampling influential points (De Bin et al 2016).

Ultimately, ESS does not uniquely focus on one resampling strategy over another, but instead uses multiple strategies to transparently assess stability both within and across forms of sampling variation.

#### 2.3.2. Sparsity Levels

In addition to the resampling strategy, the other component of the model fitting process that is varied in each analytic configuration of ESS is the sparsity level of the base learner, which can be defined as an operating characteristic that yields more or less sparse variable selection. In the case of LASSO, the operating characteristic corresponding to various sparsity levels is the penalty parameter, typically denoted λ. In this application, we define a set of penalty parameters corresponding to a specific target number of selected predictors from each model run (e.g., 5, 10, …, 25 for range and interpretability). The CPSS portion of ESS is implemented using the *stabs* package in R (Hofner et al 2015; Hofner and Hothorn 2021). This implementation takes the target number of selected predictors as input, with the LASSO penalty parameter λ adapted within each CPSS subsample to yield a model of the specified size.

In contrast, we use the R package *glmnet* for the bootstrap resampling portion of ESS (Friedman et al 2010), and the LASSO penalty parameter λ is held fixed across resamples. These parameters are calibrated in a full-dataset reference model, using the glmnet default λ sequence, to target specified model sizes. The largest λ among those yielding the same model size is selected, resulting in approximately the desired number of selected predictors within each resample.

The goal of exploring a range of sparsity levels is to capture different signal structures and to reveal how predictors behave under different levels of competition. This consideration is particularly relevant to aging biology, where candidate predictors may be correlated or associations distributed across several predictors (Cohen et al 2020, Hodgins et al 2026).

As mentioned earlier, LASSO can be sensitive to the choice of penalty parameter, so evaluating a range of sparsity levels will help to quantify the sensitivity. Traditionally, a single penalty level is chosen for a LASSO model, determined by optimizing the prediction error using cross-validation. Since selection, and not prediction, is the goal of ESS, this is not necessarily the best way to choose a sparsity level. No single sparsity level is guaranteed to capture the complexity of finite sample data, which is often small, optimally. This reflects the principle that robustness and stability in variable selection should be examined across multiple modeling conditions.

While the choice of a specific set of sparsity levels is subjective and will influence the final set of variables chosen for the grand stable set, this challenge is inherent to any hyperparameter-selection procedure. Its impact can be mitigated through sensitivity analyses or by exploring a wider or more granular range.

### 2.4. Stability Metrics and Summary Outputs

Once we complete the ESS and obtain our grand stable set, we need to condense the results into interpretable summaries. To do this, we use several stability metrics, including selection probabilities and top-*k* ranking. We also report descriptive effect sizes and directionality.

#### 2.4.1. Selection Probabilities

The selection probabilities of each predictor for each analytic configuration (i.e., combination of resampling strategy and sparsity level) can be derived directly from ESS. Although this framework is not inferential, we consider the selection probability (SP) as a measure of stability under analytic perturbation, indicating how often the predictor survives the model fitting within each configuration. We report the median SP across all configurations as a summary measure, as it is robust to outlier configurations and provides a global measure of stability for each predictor in the grand stable set.

#### 2.4.2. Top-*k* Ranking and Grand Stable Set

In addition to examining the raw selection proportions, we also consider a top-*k* ranking of variables for each configuration. This ranking is more robust than interpreting the absolute SP magnitudes alone, as SP is expected to differ across configurations. Therefore, we cannot reasonably rely on a single SP threshold to construct our stable sets, as is done in classic stability selection (Meinhausen and Buhlmann 2010, Shah and Samworth 2013). Using top-*k* rankings highlights the relative competitiveness of variables within each configuration; these rankings form the interim stable sets, and ultimately, the grand stable set.

To maximize transparency, the grand stable set is created using the union of interim stable sets. Bootstrap and CPSS emphasize different aspects of variability and varying penalization levels will yield different sparsity patterns; as such, reporting a union will allow us to collect all candidate predictors surviving at least one configuration, which is appropriate for our exploratory framework. Additionally, we annotate the grand stable set with a stability score, defined as the number of interim stable sets each variable appears. Using a union also protects against false negatives, ensuring we do not discard a real effect that may be unstable in some modes of perturbation. It may also, however, lead to a higher risk of false positives, accumulating some noise variables that survive certain perturbations by change or sample-specific characteristics. Ultimately, reporting the union gives a transparent, broad discovery set that could be pared down further depending on the downstream goals.

### 2.5. Descriptive Effect Sizes and Directionality

After obtaining the grand stable set, we perform regression models, both unadjusted and adjusted for all other predictors in the grand stable set, to quantify descriptive effect sizes and directionality of the effect for each stable predictor. No uncertainty quantification (i.e., standard errors, confidence intervals, or p-values) can be accurately calculated post-ESS due to the data-driven, ensemble-type selection approach. Instead, we use our stability metrics as a measure of confidence in robustness, and these effect measures are solely descriptive at this stage of discovery.

### 2.6. Applicability Across Predictor Domains

We motivated our framework in the context of biomarker discovery, but ESS can be applied to any type of predictor domain. Clinical predictors may not face the same problems as biomarkers (e.g., correlated signals distributed across many predictor candidates), however, they will have selection variability, effect-size instability, sensitivity to the choice of tuning parameter, and variation across resampling techniques. While analyzing a set of non-biomarker predictors may be simpler than a biomarker set, selection stability is not guaranteed using a single iteration of LASSO or another base learner.

This unified framework enables fair comparison across domains and nested groups of predictors. In our data example, we look at ESS metrics for a biomarker-only model, a clinical-only model, and a combined model with all of the above competing against each other. The creation of nested predictor sets can help determine different levels of competitiveness within and across variable types.

### 2.7. Practical Implementation Considerations

#### 2.7.1. Coding of Predictors

With respect to variable coding, continuous variables are standardized (i.e., centered and scaled) at the level of the full sample. Categorical variables are represented as dummy variables for each non-referent level of the variable. They can be input to the model using standard LASSO, where these dummy variables are independently selected as active or inactive, or input using group LASSO, wherein the dummy variables are “tethered” together and are either chosen as active or inactive as a whole variable. Neither option is inherently right or wrong; the choice depends on the scientific question. If level-specific effects are of interest, standard LASSO may be preferable. If the goal is to interpret each categorical variable as a single predictor, group LASSO may be more appropriate.

#### 2.7.2. Missing Data Considerations

Missing data are often unavoidable in high dimensional predictor sets, and ESS requires some handling of missing data before use. In particular, each LASSO run requires a complete predictor input matrix with no missing values, which can be achieved through several preprocessing strategies. ESS can accommodate multiple approaches to handling missing data, each of which may influence downstream selection probabilities, rankings, and stable sets. In the data example in Section 3, a missing indicator approach is used, where missing values for each variable are encoded as a separate variable, set to 1 if missing and 0 if observed. This approach was chosen since it preserves the full analytic sample and allows missingness patterns to be evaluated as potential predictors. Another approach is the complete case analysis, where any observations with missing data are removed. This approach may be reasonable when the percentage of missing data is small and missingness is conditionally independent of the outcome. Single or multiple imputation are common in applied data analyses as well. Single imputation requires one imputation for each missing observation. Single imputation is straightforward to implement but the imputed value is treated as if it were observed; this ignores uncertainty associated with the imputation process, which can lead to downstream variable-selection results that are sensitive to the imputation technique used. Multiple imputation is often regarded as a preferred approach in applied settings, but it can quickly become computationally intensive with ESS, since the entire pipeline would need to be run on each imputed dataset, and an approach for combining selection probabilities, rankings, and stable sets across imputations would need to be specified. Ultimately, because the goal of ESS is to identify stable predictor sets rather than estimate population parameters, the operationalization of missingness should be guided by the intended scientific or clinical application. The results of ESS may be sensitive to the choice of missing data handling technique, so sensitivity analyses are recommended.

#### 2.7.3. Guidance for Parameter Choices

In order to use ESS, the user must choose their set of sparsity levels, the number of resamples per analytic configuration, *B*, and the number of top predictors to include in each interim stable set, *k*. The parameter choices are context-dependent and should be explored but are not optimized for a particular performance measure. The results of ESS may be sensitive to the choice of these parameters, so additional sensitivity analyses varying these parameters are recommended. However, not all parameters are expected to contribute equally to variation in results; sensitivity will likely be greatest for the sparsity levels, which directly governs predictor competition, followed by the choice of *k*, which affects the breadth of the interim stable sets.

In choosing the set of sparsity levels, it is important to consider any background knowledge of the predictor set, the size of the predictor set, and the size of the sample. Background knowledge of the predictor set can provide some guidance regarding minimum or maximum number of active predictors in the set. To avoid oversaturating the corresponding model in our data analysis example, we choose a minimum active predictor set size of 5 due to our anticipated number of active predictors in the set, and a maximum of 25, representing approximately a quarter of the smallest sample size we were using. The coarseness of the sparsity level set will affect computational speed; we chose steps of five.

The implementation parameters *B* and *k* control the resolution of stability assessment. Computational speed will be a consideration when choosing the number of resampling iterations per configuration. For stability selection, *B* = 100 is recommended as sufficient (Meinhausen and Buhlmann 2010, Shah and Samworth 2013), and for Bolasso, increasing *B* is always appears beneficial (Bach 2008). In our illustrative data analysis example, we choose *B* = 200 as a compromise between these recommendations. The choice of *k* in the top-*k* rankings is application-dependent; in our illustrative data analysis example, we chose to keep the top 20% of the candidate predictors in the interim stable set for each configuration. This specific choice may be infeasible and uninformative for extremely high-dimensional predictor sets or other application contexts.

#### 2.7.4. Downstream Prioritization and Interpretation of Stable Sets

As discussed, the grand stable set is built with the union of the interim stable sets for maximum transparency and broad discovery. If the goal is to gather a minimal set of highly reliable variables, the intersection of the interim stable sets could be used alternatively to prioritize predictors with the most consistency across configurations. This would highlight the candidate covariates that survive the repeated reweighting of observations from bootstrap resampling, the structured perturbations from CPSS, and the changing sparsity levels that allow for various degrees of predictor competition. This set may have a higher false negative rate but may indicate the highest priority candidates for validation.

Alternatively, it might be useful to combine aspects of the union and intersection approaches. For example, one might retain variables appearing in at least three interim stable sets, variables present in both resampling modes at least once, or variables recurring across several sparsity levels. These prioritization strategies represent downstream choices that can be made after running ESS, depending on the goals of the analysis.

## 3. Data Example

### 3.1. Study Design and Cohort Description

To illustrate the utility of ESS, we provide a real-data example using the motivating research topic of physical resilience in older adults. The Physical Resilience Indicators and Mechanisms in the Elderly (i.e., PRIME) Collaborative has the goal of identifying predictors of resilience. The current study of interest, PRIME-KNEE (PK), examines physical resilience following elective total knee arthroplasty. The methods related to PK have been published previously (Whitson et al 2021). Briefly, PK enrolled 203 community-dwelling adults over the age of 60 years who were scheduled to undergo unilateral total knee arthroplasty (TKA). An extensive set of measures was collected pre-surgery (at baseline), including measures of physical, cognitive, and psychosocial reserve, provocative tests, demographics, comorbidities, blood biomarkers, and resilience health measures. The same resilience health measures were collected longitudinally for 6 months post-surgery; these included PROMIS (Patient-Reported Outcomes Measurement Information System) pain intensity and interference scales, daily step counts recorded from Garmin devices, lower extremity activities of daily living scale (LE PADL), attention items of the 3-Minute Diagnostic Confusion Assessment Method (3D-CAM), and cognitive change index (CCI). Although these health measures span multiple domains, including cognition, we focus on physical resilience outcomes in PK analyses because their recovery trajectories exhibited more meaningful heterogeneity, whereas the cognitive outcomes (i.e., 3D-CAM and CCI) demonstrated early ceiling effects in this cohort.

### 3.2. Brief Overview of Measuring Physical Resilience Profiles

Resilience following a stressor is inherently difficult to quantify; our previous work has introduced several outcome measures to operationalize the concept of resilience, including the previously described Expected Recovery Differential (ERD) and the Recovery Trajectory (RT) approaches (Colón-Emeric et al 2020). In our data example, we use an outcome derived from the RT classes for PRIME-KNEE. Colón-Emeric et al (2026) describe the determination of trajectory classes for the PK cohort. Briefly, latent class trajectory analysis was used to group resilience outcome trajectories over the recovery period into a finite number of classes. Then, each participant was given a probability of belonging to each class based on their observed trajectory. In this analysis, we use a continuous version of the RT classes for our model; namely, our resilience outcome is the probability of belonging to a highly resilient trajectory class.

### 3.3. Predictor Sets and ESS Specifications

Various analyses of the PK data are ongoing (e.g., Colón-Emeric et al 2026). Here, we show an illustrative example using ESS that is not being published elsewhere. Specifically, we focus on the probability of belonging to a highly resilient trajectory class for the PROMIS pain interference health measure. The outcome is coded such that higher values indicate more resilience, to align with intuitive expectation of a resilience outcome. We applied ESS to three candidate predictor sets: clinical predictors, plasma blood biomarkers, and a combination of the two. Predictor sets were separated into clinically accessible variables and plasma biomarkers to facilitate downstream evaluation of whether routinely available measures provide sufficient information for identifying resilience-associated signals or whether additional information is gained from specialized biomarker assays.

The clinical predictor set is comprised of a set of variables we consider ‘clinically accessible’, including baseline demographics, comorbidities, physical, cognitive, and psychosocial reserve measures, and laboratory measures like eGFR (full list and details in Supplemental Table 1). Predictors were encoded using standard LASSO, rather than group LASSO, with missing data accounted for using a missing-indicator approach. For continuous variables, a separate indicator encoded missingness while missing values in the original predictor were set equal to the sample mean. Meanwhile, for categorical variables, missingness was simply encoded using a separate indicator for missingness. This resulted in 48 total candidate predictors for the clinical set. The biomarker predictor set consisted of immune biomarkers, quantified in plasma, that are typically not considered ‘clinically accessible’ markers. This set had 49 plasma biomarkers, including one semi-continuous marker, CMVIgG, coded using an absence indicator and a continuous concentration among positive values (full list and details in Supplemental Table 2). The combined predictor set consisted of all 97 predictors.

We used both the bootstrap and CPSS methods of resampling, and we chose 5 penalty parameters for LASSO corresponding to approximately 5, 10, 15, 20, and 25 active predictors in the model. This leads to 10 total analytic configurations. Additionally, we chose *B* = 200 to be the number of resamples per analytic configuration and *k* = 10 predictors in each interim stable set. Finally, when summarizing descriptive effect sizes and directionality for predictors in the grand stable sets, adjusted regression models were controlled for all other stable predictors as well as a post-operative illness variable, which indicates self-reported injury/illness that prevented performance of everyday activities for at least one day during the recovery period. All analyses were completed using R 4.4.0 (R Core Team 2024). The code used to implement ESS for this data example is publicly available on Github (see Data Availability Statement).

### 3.4. Results

Table 1 includes select descriptive characteristics of the analytic sample. Of the 203 participants enrolled in PRIME-KNEE, 4 did not undergo knee replacement surgery and therefore had no post-surgery pain interference measurements. An additional 49 participants did not have baseline plasma biomarkers available, yielding a final analytic sample of 150 participants. The sample was predominantly white and non-Hispanic/Latino, with a higher proportion of female participants and relatively low financial stress. The average age at surgery is 71.4 years (SD 6.5), and the average education level is 16.1 years (SD 2.3).

**Table 1:** Descriptive Characteristics of the Analytic Sample. *Values are n (%) unless otherwise noted*.

|  |  |
| --- | --- |
|  | Overall (N=150) |
| **Age at Surgery Date (years)** |  |
| Mean (SD) | 71.4 (6.48) |
| Missing | 2 (1.3%) |
| **Race** |  |
| White | 126 (84.0%) |
| Black or African American | 19 (12.7%) |
| Asian | 2 (1.3%) |
| More than one race | 2 (1.3%) |
| Unknown or Not Reported | 1 (0.7%) |
| **Ethnicity** |  |
| Not Hispanic or Latino | 145 (96.7%) |
| Hispanic or Latino | 4 (2.7%) |
| Unknown or Not Reported | 1 (0.7%) |
| **Sex** |  |
| Female | 92 (61.3%) |
| Male | 58 (38.7%) |
| **Education Level (years)** |  |
| Mean (SD) | 16.1 (2.31) |
| Missing | 1 (0.7%) |
| **Financial Stress** |  |
| After paying the bills, you still have enough money for special things that you want | 116 (77.3%) |
| You have enough money to pay the bills, but little spare money to buy special things | 21 (14.0%) |
| You have money to pay the bills, but only because you have to cut back on things | 3 (2.0%) |
| You are having difficulty paying the bills, no matter what you do | 1 (0.7%) |
| Missing | 9 (6.0%) |

Table 2 includes results from the grand stable set in the clinical predictors-only for the clinical predictors that remained in the grand stable set of the combined model. PHQ-9 (Patient Health Questionnaire-9), which indicates depressive symptoms, emerged as the most robust predictor in both the clinical model and the combined model, based on a perfect 10/10 stability score and high median selection probability across analytic configurations. Higher depressive symptoms were associated with a lower probability of being in a highly resilient trajectory class for pain interference, which directionally aligns with prior expectations.

**Table 2:** Grand Stable Set for Clinical Only and Combined Models.

| Predictor | Description | Clinical Only Model |  |  |  | Combined Model* |  |  |
| --- | --- | --- | --- | --- | --- | --- | --- | --- |
|  |  | Unadj Coef | Adj Coef | Stability Score | Median SP | Adj Coef | Stability Score | Median SP |
| PHQ9<sup>†</sup> | Higher score -> More Depression | -0.122 | -0.126 | 10 | 0.958 | -0.109 | 10 | 0.835 |
| Diabetes: No | n = 110 | -0.152 | -0.227 | 10 | 0.948 | -0.203 | 10 | 0.780 |
| Race: Black/AA | n = 19 | -0.294 | -0.234 | 10 | 0.927 | -0.221 | 10 | 0.785 |
| Trail-Making Test B: Missing | n = 6 | -0.515 | -0.428 | 10 | 0.867 | -0.320 | 10 | 0.772 |
| Race: More than 1 | n = 2 | -0.528 | -0.756 | 10 | 0.732 | -0.634 | 9 | 0.593 |
| Financial Stress: Enough \$ but Little Spare | n = 21 | -0.199 | -0.107 | 10 | 0.655 | -0.118 | 7 | 0.455 |
| Age | Higher value -> Older | 0.049 | 0.032 | 10 | 0.580 |  |  |  |
| Financial Stress: Difficulty w/ Bills | n = 1 | -0.534 | -0.336 | 4 | 0.477 |  |  |  |
| Race: Missing | n = 1 | -0.530 | -0.578 | 4 | 0.475 |  |  |  |
| Race: Asian | n = 2 | -0.028 | -0.035 | 3 | 0.473 |  |  |  |
| 3-Minute Walk | Higher score -> Better Walking Endurance | 0.049 | 0.030 | 3 | 0.468 |  |  |  |
| Ethnicity: Missing | n = 1 | 0.473 | 0.351 | 3 | 0.465 |  |  |  |
| 3MS<sup>†</sup> | Higher score -> Better Cognitive Functioning | 0.056 | 0.029 | 3 | 0.463 |  |  |  |
| Trail-Making Test A | Higher score -> Slow Processing | -0.066 | 0.022 | 3 | 0.258 | 0.001 | 1 | 0.220 |
| Psychosocial Resilience Scale: Missing | n = 30 | -0.106 | -0.009 | 2 | 0.465 |  |  |  |
| 15-Item Word List | Higher score -> Better Verbal Memory | 0.031 | 0.030 | 2 | 0.463 |  |  |  |
| 3MS<sup>†</sup> Missing | n = 4 | -0.258 | -0.115 | 2 | 0.417 |  |  |  |
| eGFR<sup>†</sup> | Higher score -> Better Kidney Function | 0.006 | 0.033 | 1 | 0.392 |  |  |  |
| eGFR<sup>†</sup>: Missing | n = 8 | 0.042 | 0.084 | 1 | 0.350 |  |  |  |

\*Blank entries indicate predictors that did not appear in the grand stable set for the combined model.
<sup>†</sup> PHQ9: Patient Health Questionnaire-9; 3MS: Modified Mini-Mental State Test; estimated Glomerular Filtration Rate

Most clinical predictors with perfect stability scores in the clinical-only model remained competitive in the combined model, indicating robustness even in the presence of the plasma blood biomarkers. Diabetes and race are notable examples. Diabetes (No vs Yes) was negatively associated with probability of a highly resilient trajectory, which is an unexpected direction worth evaluating further in other cohorts.

Black/African American race (vs White) was also negatively associated with probability of a highly resilient trajectory. Additionally, several missingness indicators appeared in the grand stable set, indicating that missingness patterns may carry relevant information regarding resilience outcomes. The interpretation of these effects is unclear, especially for the indicators with low prevalence; they should be viewed as descriptive and are simply reported as part of the exploratory process.

Table 3 is analogous to Table 2 for the biomarker predictor set. Here, we again observe that the most robust biomarkers from the biomarker-only model are also robust when competing against the clinical predictors in the combined model. In this case, the level of robustness varied depending on which predictors they were competing against. IL27, IL7, Leptin, and IL1Ra were very stable and highly selected in both models. LBP (Lipopolysaccharide-Binding Protein) and IL5 were much more robust to analytic configuration when competing only against biomarkers, which suggests that LBP and IL5 may share signal with clinical predictors.

**Table 3:** Grand Stable Set for the Biomarker Only and Combined Models.

| Predictor† | Biomarker Only Model |  |  |  | Combined Model* |  |  |
| --- | --- | --- | --- | --- | --- | --- | --- |
|  | Unadj Coef | Adj Coef | Stability Score | Median SP | Adj Coef | Stability Score | Median SP |
| IL27 | 0.091 | 0.095 | 10 | 0.930 | 0.056 | 10 | 0.665 |
| IL7 | 0.076 | 0.075 | 10 | 0.837 | 0.063 | 10 | 0.650 |
| LBP | 0.055 | 0.060 | 10 | 0.782 | 0.041 | 2 | 0.353 |
| IL5 | -0.048 | -0.072 | 10 | 0.677 | -0.034 | 3 | 0.387 |
| Leptin | -0.066 | -0.054 | 10 | 0.657 | -0.028 | 10 | 0.420 |
| IL1Ra | -0.061 | -0.045 | 9 | 0.532 | -0.050 | 9 | 0.448 |
| IL37 | 0.055 | 0.034 | 9 | 0.498 |  |  |  |
| LIF | 0.053 | 0.040 | 8 | 0.552 |  |  |  |
| IL15 | -0.038 | -0.059 | 6 | 0.512 |  |  |  |
| IFNa2a | 0.041 | 0.031 | 6 | 0.490 |  |  |  |
| IFNy | 0.031 | 0.033 | 5 | 0.485 |  |  |  |
| IL1B | -0.035 | -0.037 | 4 | 0.458 |  |  |  |
| TNFRI | -0.036 | -0.058 | 1 | 0.442 |  |  |  |
| IL8 | -0.030 | -0.016 | 1 | 0.425 |  |  |  |
| Cortisol | 0.043 | 0.005 | 1 | 0.365 |  |  |  |
| TNFa | 0.016 | 0.070 | 1 | 0.280 |  |  |  |

\*Blank entries indicate predictors that did not appear in the grand stable set for the combined model.
† More information on each biomarker can be found in Supplemental Table 2.

Supplemental Figure 1 complements the tables with configuration-level heat maps to help further explain the robustness results. This figure shows exactly which predictors entered the interim stable sets under each resampling strategy and sparsity level combination, and they also provide a gradient indicating the selection probabilities. For example, Trail-Making Test A appeared in limited configurations in the clinical-only model and remained competitive for only one configuration in the combined model. The heatmaps show that Trail-Making Test A was relatively competitive in the configurations with higher sparsity levels suggesting sensitivity to predictor competition. We similarly explored configuration-level results for IL5 and LBP; these biomarkers showed reduced competitiveness in the combined model, persisting only under lower sparsity levels.

Overall, the analyses across clinical-only, biomarker-only and combined predictor domains revealed many meaningful patterns. Some predictors were consistently stable across analytic configurations and predictor-sets, while others were competitive only in certain contexts dependent on the sparsity level, resampling strategy, and the other candidate predictors.

## 4. Discussion

The results from the data example demonstrate how ESS characterizes the stability of candidate predictors across different data perturbations and sparsity levels. The comparison across predictor domains further shows the robustness of signals across different levels of predictor competition. The implications of these findings, methodological limitations and further considerations are discussed below.

### 4.1. Implications of the Data Example and Downstream Use of ESS Outputs

The PRIME-KNEE data example illustrates the exploratory and transparent nature of ESS. In this application, the pain interference recovery trajectory revealed one particularly stable clinical predictor, PHQ-9, exhibiting superior stability scores and selection probabilities across both the clinical-only and combined predictor sets. Diabetes also showed consistently high stability metrics, although the direction of association was counter to expectations. It is unclear whether this is an artifact of the highly selective PK sample (i.e., selected for elective surgery) or reflects a true generalizable signal. The signal may be related to consistency of medical care or other unmeasured confounders, such as the use of metformin (i.e., a medication under study as a gerotheraputic). Further exploration in independent cohorts is warranted. In addition, the consistent appearance of race (Black/African American vs. White) and financial stress indicators underscores the potential importance of socioeconomic context in resilience research and highlights the need for broader representation and more granular data in future studies. ESS, by design, reveals these stable signals that can inform prioritization for downstream validation. An important next step is evaluation of stable candidate predictors in independent cohorts to determine the extent to which they generalize across study populations and resilience outcome definitions. These validation efforts can help distinguish between predictors that are associated with resilience consistently across settings versus those that are stable in a particular cohort.

ESS also identified several stable biomarkers that appear competitive, even against commonly measured clinical predictors. Some biomarkers are less accessible for routine clinic use, so it may be a priority to determine if the addition of those biomarkers can predict or characterize resilience outcomes beyond clinical variables alone. One potential downstream use of ESS outputs is to explore predictive performance across domains using models derived from grand stable sets, or selected subsets, of each predictor set. This should be done cautiously, as the prediction models are run on the same dataset used for ESS, and any findings should therefore be interpreted as hypothesis-generating.

Broadly, ESS outputs can be used to prioritize candidate variables to be incorporated, highlighted, or validated in independent studies or for further exploratory analyses within the analytic sample.

### 4.2. Methodological Limitations and Considerations

ESS also has limitations as well as design considerations. Although ESS is inspired by stability selection, which gives some theoretical guarantees related to error control, ESS itself does not claim these guarantees, since it is an ensemble approach incorporating multiple types of resampling and sparsity levels. Additionally, although we report descriptive effect sizes and directionality, we cannot quantify the uncertainty related to these estimates, because ESS lies far outside the assumptions underlying existing post-selection inference framework. That said, inference is not the primary aim of ESS; its purpose is exploratory variable selection. This should be made explicit whenever ESS results are presented, as the goal is not to confirm any associations or predictor-outcome relationships.

Like any variable selection technique, ESS may be sensitive to design choices such as the base learner, resampling schemes, sparsity levels, and top-*k* size. In the methods and real data example, we described some potential means of selecting these values, but they are not optimized for any prediction or inference. Therefore, we recommend pre-specifying these design choices, reporting them transparently, and considering sensitivity analyses. Also related to our data example, the ESS outputs are limited due to the lack of interpretation for the missingness-indicator approach. Especially for variable-levels with small sample sizes, which are particularly prone to highly influential points, the observed stable signals may be driven by artifacts of the analytic sample. This issue also underscores the broader limitation of sample size, which similarly affects generalizability.

Although ESS can be applied across a range of sample sizes and predictor dimensions, formal sample size requirements or theoretical guarantees regarding recovery of a true predictor set have not yet been established. Like other resampling-based variable selection approaches, larger sample sizes are expected to yield more stable selection probabilities because resampled datasets are more likely to capture the underlying signal structure and better distinguish among competing predictors. Conversely, as the number of candidate predictors increases, so does predictor competition, potentially leading to greater sensitivity to sparsity and implementation choices. Future methodological work, including simulation studies, is needed to characterize the performance of ESS across different sample size and predictor-dimension settings, as well as signal strengths and correlation structure among the predictors.

### 4.3. Conclusions

In this paper, we have proposed an Exploratory Stability Selection (ESS) framework, a variable selection pipeline that emphasizes robustness, consistency, and transparency in hypothesis-generating predictor discovery, rather than focusing on single model results. ESS is well-suited for correlated, lower signal predictors as well as mixed-type predictors, all of which are common in aging research and especially in discovery of factors related to physical resilience. Practically, we recommend using ESS for hypothesis generation and exploratory screening when there are many candidate predictors of interest, followed by structured validation or confirmation studies where appropriate.

## CRediT Authorship Contribution Statement

**Marissa C. Ashner**: Conceptualization, Methodology, Software, Formal Analysis, Data Curation, Writing – Original Draft, Visualization. **Virginia B. Kraus:** Conceptualization, Resources, Writing – Review & Editing, Supervision. **Heather E. Whitson**: Conceptualization, Writing – Review & Editing, Supervision, Funding Acquisition. **Corey B. Simon**: Conceptualization, Writing – Review & Editing. **Janet L. Huebner:** Conceptualization, Resources, Writing – Review & Editing. **Akshay Bareja**: Conceptualization., Software, Writing – Review & Editing, Visualization. **Chelsea R. Perfect**: Conceptualization, Writing – Review & Editing. **Laura Pietrosimone**: Conceptualization, Writing – Review & Editing. **Katherine S. Hall**: Conceptualization, Writing – Review & Editing. **Cathleen S. Colón-Emeric:** Conceptualization, Writing – Review & Editing, Supervision, Funding Acquisition. **Sarah B. Peskoe**: Conceptualization, Methodology, Formal Analysis, Writing – Review & Editing, Supervision.

## Declaration of generative AI and AI-assisted technologies in the manuscript preparation process

During the preparation of this work, the authors used Microsoft Copilot to assist with editing, language refinement, and organization of manuscript text. After using this tool, the authors reviewed and edited the content as needed and take full responsibility for the content of the published article.

## Data Availability

Data from this study are not publicly available, but de-identified datasets can be requested via email to the corresponding author. The R code used to implement the ESS framework is publicly available on Github at https://github.com/marissaashner/ExploratoryStabilitySelection.

