## Supplemental Material for "An Exploratory Stability Selection (ESS) Framework for Robust Predictor Discovery: An Application to Physical Resilience in Aging Populations"

**Supplemental Table 1: Candidate Predictors in the Clinical-Only ESS Model taken at baseline**

| **Variable** | **Description / Levels** | **Frequency (%) Missing** |
| --- | --- | --- |
| Age at Surgery | Continuous (in years) if data available, plus a missing indicator | 2 (1.33%) |
| Biological Sex | Categorical (Ref: Female / Male) | 0 (0%) |
| Self-reported Race | Categorical (Ref: White / Black / Asian / More than One / Missing) | 1 (0.67%) |
| Self-reported Ethnicity | Categorical (Ref: Not Hispanic or Latino / Hispanic or Latino / Missing) | 1 (0.67%) |
| Financial Stress | Categorical (Enough: Ref / Little to Spare / Need to Cut Back / Have Difficulty / Missing) | 9 (6.00%) |
| Education | Continuous (in years completed) if data available, plus a missing indicator | 1 (0.67%) |
| Vascular Disease | Categorical (Yes: Ref / No / Missing) | 6 (4.00%) |
| Heart Disease | Categorical (Yes: Ref / No / Missing) | 6 (4.00%) |
| Chronic Liver Disease | Categorical (Yes: Ref / No / Missing) | 7 (4.67%) |
| Diabetes | Categorical (Yes: Ref / No / Missing) | 6 (4.00%) |
| History of treated non-skin cancer | Categorical (Yes: Ref / No / Missing) | 6 (4.00%) |
| eGFR (estimated Glomerular Filtration Rate) | Continuous if data available, plus a missing indicator | 8 (5.33%) |
| BMI (Body Mass Index) | Continuous (in kg/m^2) if data available, plus a missing indicator | 2 (1.33%) |
| Gait Speed | Continuous (in m/sec) | 0 (0%) |
| 3-minute walk | Continuous (in feet) | 0 (0%) |
| grip strength | Continuous (in kg) | 0 (0%) |
| 3MS (Modified Mini-Mental State Test) | Continuous if data available, plus a missing indicator | 4 (2.67%) |
| Trail Making Test B | Continuous (in sec) if data available, plus a missing indicator | 6 (4.00%) |
| Trail Making Test A | Continuous (in sec) if data available, plus a missing indicator | 1 (0.67%) |
| 15 item word list | Continuous (number of items recalled) if data available, plus a missing indicator | 1 (0.67%) |
| Digit Symbol Substitution Test | Continuous (number completed) if data available, plus a missing indicator | 4 (2.67%) |
| Resilience Scale | Continuous if data available, plus a missing indicator | 30 (20.0%) |
| PHQ-9 (Patient Health Questionnarie-9) | Continuous if data available, plus a missing indicator | 15 (10.0%) |
| PROMIS SF (Patient-Reported Outcomes Measurement Information System Short Form) Emotional Support 4a | Continuous (t-score) if data available, plus a missing indicator | 8 (5.33%) |

| **Biomarker (Plasma)** | **Molecular Class** | **Primary Function** |
| --- | --- | --- |
| Activin A | Protein | Cell differentiation |
| C-Reactive Protein (CRP) | Protein | Acute phase inflammation |
| Cortisol | Hormone | Stress response |
| Cytomegalovirus-Specific IgG (CMVIgG) | Antibody | CMV infection (Concentration) |
| CMVIgG Negative |  | CMV infection (Yes: Ref / No) |
| D-Dimer | Protein | Clot breakdown marker |
| Eotaxin (CCL11) | Chemokine | Eosinophil recruitment |
| Fibrinogen | Protein | Coagulation Factor |
| Glycoprotein 130 (gp130) | Protein | IL-6 signaling |
| Granzyme B (GZMB) | Protein | Activate inflammatory cytokines |
| Growth-Regulated Oncogene Alpha (GRO-α, CXCL1) | Chemokine | Neutrophil recruitment |
| Growth Differentiation Factor 15 (GDF-15) | Protein | Stress response |
| Interferon-Inducible T-cell-α Chemoattractant (I-TAC, CXCL11) | Chemokine | T-cell recruitment |
| Intercellular Adhesion Molecule-1 (ICAM-1) | Protein | Leukocyte adhesion |
| Interferon Alpha 2a (IFN-α2a) | Cytokine | Antiviral response |
| Interferon Gamma (IFN-γ) | Cytokine | Macrophage activation |
| Interferon Gamma-Induced Protein 10 (IP-10, CXCL10) | Chemokine | T-cell recruitment |
| Interleukin-1 Beta (IL-1β) | Cytokine | Proinflammatory mediator |
| Interleukin-1 Receptor Antagonist (IL-1Ra) | Cytokine | IL-1 inhibition |
| Interleukin-2 (IL-2) | Cytokine | T-cell proliferation |
| Interleukin-4 (IL-4) | Cytokine | B-cell differentiation |
| Interleukin-5 (IL-5) | Cytokine | Eosinophil activation |
| Interleukin-6 (IL-6) | Cytokine | Acute-phase response |
| Interleukin-7 (IL-7) | Cytokine | T-cell development |
| Interleukin-8 (IL-8, CXCL8) | Chemokine | Neutrophil recruitment |
| Interleukin-10 (IL-10) | Cytokine | Anti-inflammatory |
| Interleukin-12p70 (IL-12p70) | Cytokine | Th1 differentiation |
| Interleukin-13 (IL-13) | Cytokine | Allergic response |
| Interleukin-15 (IL-15) | Cytokine | NK cell survival |
| Interleukin-18 (IL-18) | Cytokine | IFN-γ induction |
| Interleukin-27 (IL-27) | Cytokine | Immune regulation |
| Interleukin-37 (IL-37) | Cytokine | Anti-inflammatory |
| Leptin | Hormone | Energy homeostasis |
| Leukemia Inhibitory Factor (LIF) | Cytokine | Cell differentiation |
| Lipopolysaccharide (LPS) | Protein | Innate immunity trigger |
| Lipopolysaccharide-Binding Protein (LBP) | Protein | LPS transport/innate immunity |
| Monocyte Chemoattractant Protein-1 (MCP-1, CCL2) | Chemokine | Monocyte migration |
| Monokine Induced by Gamma Interferon (MIG, CXCL9) | Chemokine | T-cell recruitment |
| Macrophage Inflammatory Protein-1 Alpha (MIP-1α, CCL3) | Chemokine | Granulocyte activation |
| Macrophage Inflammatory Protein-1 Beta (MIP-1-β, CCL4) | Chemokine | Leukocyte trafficking |
| Plasminogen Activator Inhibitor-1 (PAI-1) | Protein | Fibrinolysis inhibition |
| Serum Amyloid A (SAA) | Protein | Acute-phase inflammation |
| Tumor Necrosis Factor Receptor Superfamily Member 8 (TNFRSF8, CD30) | Protein | Immune activation |
| TNF-Related Apoptosis-Inducing Ligand (TRAIL) | Protein | Induces apoptosis |
| Transforming Growth Factor Alpha (TGF-α) | Protein | Cell proliferation |
| Transforming Growth Factor Beta (TGF-β) | Cytokine | Immune regulation |
| Tumor Necrosis Factor Alpha (TNF-α) | Cytokine | Systemic inflammation |
| Tumor Necrosis Factor Receptor I (TNFRI) | Protein | TNF signaling |
| Vascular Cell Adhesion Molecule-1 (VCAM-1) | Protein | Leukocyte adhesion |

**Supplemental Table 2: Candidate Predictors in the Biomarker-Only ESS Model**

**Supplemental Figure 1: Heatmaps representing selection probabilities for predictors in the grand stable set from each predictor set. Each column represents an analytic configuration, with “B” denoting bootstrap and “C” denoting complementary pairs. The number after each letter corresponds to the approximate number of predictors retained by LASSO. Cell color reflects selection probability (SP), and yellow dots indicate selection into the top-10 ranked predictors per configuration.**

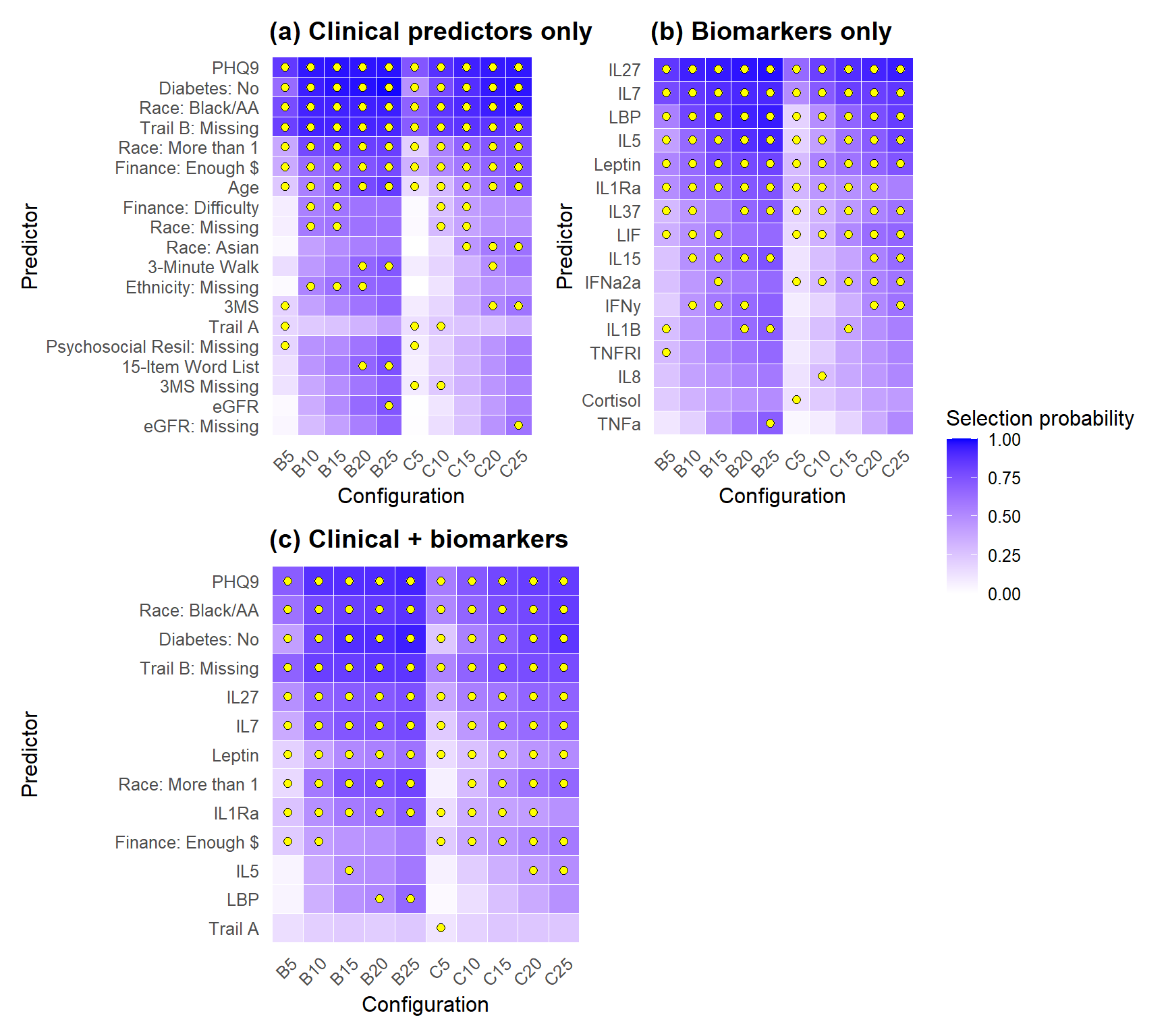
